# Charting the Decline of the Fourth Wave: US Overdose Deaths by Race, Ethnicity, and Substance Involvement

**DOI:** 10.64898/2026.01.25.26344769

**Authors:** Joseph R. Friedman, Joseph J. Palamar, Daniel Ciccarone, Tommi L. Gaines, Annick Borquez, Chelsea L. Shover, Steffanie A. Strathdee

**Author notes:** Correspondence to: Joseph Friedman, MD PhD, MPH, University of California, San Diego, 200 W Arbor Dr, San Diego, CA, 92103.

## Abstract

**Aims:** To characterize decreases in overdose death rates in the United States between 2023 and 2024 by race/ethnicity, and substance involvement.

**Design:** Population-based study of national death records accessed via the Centers for Disease Control and Prevention (CDC) Wide-ranging ONline Data for Epidemiologic Research (WONDER) platform using an underlying cause of death approach.

**Setting:** United States (US).

**Participants/cases:** All individuals who died from drug overdose between January 1999 and December 2024.

**Measurements:** Annual overdose deaths per 100,000 population. Year of occurrence of overdose death; substance involvement; race/ethnicity of decedents.

**Findings:** After many years of increases, the US overdose death rate dropped 24.4% between 2023 and 2024. Decreases reflected declining illicit fentanyl-involved deaths (with and without stimulant involvement). The fourth wave of the US overdose crisis—defined by deaths involving fentanyl together with stimulants—declined for the first time in 2024. Despite overall decreases, deaths involving stimulants without fentanyl, and xylazine, continued to represent a growing fraction of overdose fatalities. Non-Hispanic Black and African Americans had the largest decrease in death rates in 2023-2024—falling 29.3%, but remained elevated at 36.0 per 100,000, 1.51 times higher than the national average of 23.7 per 100,000. Non-Hispanic American Indian and Alaska Native individuals had the highest overdose death rates rate in 2024, at 50.8 per 100,000, representing 2.15 times the national average rate, and experienced a below-average relative decrease of 20.1%.

**Conclusions:** All four previously defined waves of the US overdose crisis are in decline, as deaths involving illicit fentanyl, with and without stimulants, are dropping sharply. Concurrently, the fraction of overdose deaths involving stimulants without fentanyl and those involving xylazine, continued to increase. While racial disparities in drug overdose death rates narrowed slightly during this period, significant gaps remain, with the highest rates among American Indian, Alaska Native, and Black individuals.

## Introduction

After many years of rapid increases, overdose death rates in the US have begun decreasing, drawing considerable public attention (1–3).

The US overdose crisis has been characterized by a ‘four-wave’ paradigm, wherein an escalating death rate was attributed to shifting drug profiles over time (4–6). The first wave entailed a surge in deaths involving commonly prescribed opioids (7). Subsequently, in wave two, drug overdose deaths involving heroin increased rapidly (8). Wave three saw the market takeover of illicitly-manufactured synthetic opioids, primarily fentanyls (8). The fourth wave quickly followed, involving the polysubstance use of fentanyl with illicit stimulants such as cocaine and methamphetamine (4,5). Deaths involving the combination of fentanyl with the veterinary sedative xylazine also increased during this period (9,10). Polysubstance overdose deaths are not a new phenomenon—for instance, benzodiazepines were frequently co-involved in prescription opioid deaths during the first wave (11)—however the fourth wave has been characterized by a particularly lethal rise in intentional and unintentional polysubstance drug use (12).

These four waves of the overdose crisis have also seen epidemiological differences by race/ethnicity. Black and African Americans have been disproportionately affected by waves three and four of the crisis (13). American Indian and Alaska Native individuals have been disproportionately affected by all waves of the overdose crisis, with disparities worsening sharply in recent years (13). One study found that death rates decreased among Non-Hispanic White individuals as of 2023, but they were still increasing among the most affected racial/ethnic minority groups, suggesting widening disparities despite overall gains at the population level (3).

Here, we leverage recent records to describe updated overdose death rates between January 1999-December 2024 by substance-involvement and race/ethnicity.

## Methods

Comprehensive national death data, representing all registered overdose deaths in the US, were obtained for the 1999-2024 period, stratified by year, month, race/ethnicity, and substance-involvement via the Centers for Disease Control and Prevention (CDC) Wide-ranging ONline Data for Epidemiologic Research (WONDER) platform using an underlying causes of death (UCD) approach (14). Overdose deaths were defined by ICD-10 UCD X40–X44 (unintentional), X60–X64 (suicide), X85 (homicide), and Y10–Y14 (undetermined intent) (14). Substance involvement was secondarily characterized using ICD-10 multiple cause of death codes, including prescription opioids (T40.2), heroin (T40.1), synthetic opioids (primarily and herein referred to as fentanyl) (T40.4) (5), cocaine (T40.5), psychostimulants with abuse potential (primarily and herein referred to as methamphetamine) (T43.6) (15), and xylazine (T42.7 or T46.5) (9).

We calculated fentanyl-deleted death rates for prescription opioids, heroin, cocaine, methamphetamine, and stimulants (either cocaine or methamphetamine), to assess trends for each substance, removing the influence of fentanyl co-involvement. For each, this entailed subtracting the death rate from the substance together with fentanyl from the total death rate for the substance.

Recorded deaths from 2024 were provisional and may slightly underestimate finalized records, whereas records from 2023 and prior were finalized. All death rates were crude per 100,000 population. All analyses were conducted in R version 4.4.1.

## Results

Figure 1 provides a simplified schematic representing trends in the four waves of the US overdose crisis from 1999 through 2024. Death rates involving prescription opioids and heroin (with fentanyl co-involved deaths omitted) reached inflection points and began to fall in 2017 and 2016, respectively, marking the decline of waves one and two of the crisis. Overdose death rates involving illicit fentanyl without stimulant co-involvement began to fall in 2022, marking the decline of wave three. Overdose death rates involving fentanyl with stimulants declined for the first time in 2024, beginning the decline of wave four. Deaths involving methamphetamine (without fentanyl co-involvement) and cocaine (without fentanyl co-involvement) continued increasing between 2023 and 2024, consistent with an overall trend of increasing stimulant-related deaths. Deaths involving xylazine also held steady and represented an increasing proportion of fentanyl-involved deaths between 2023 and 2024, rising from being co-involved in 8.3% of fentanyl-involved overdose deaths in 2023 to 12.6% of fentanyl-involved deaths in 2024.

**Figure 1.**
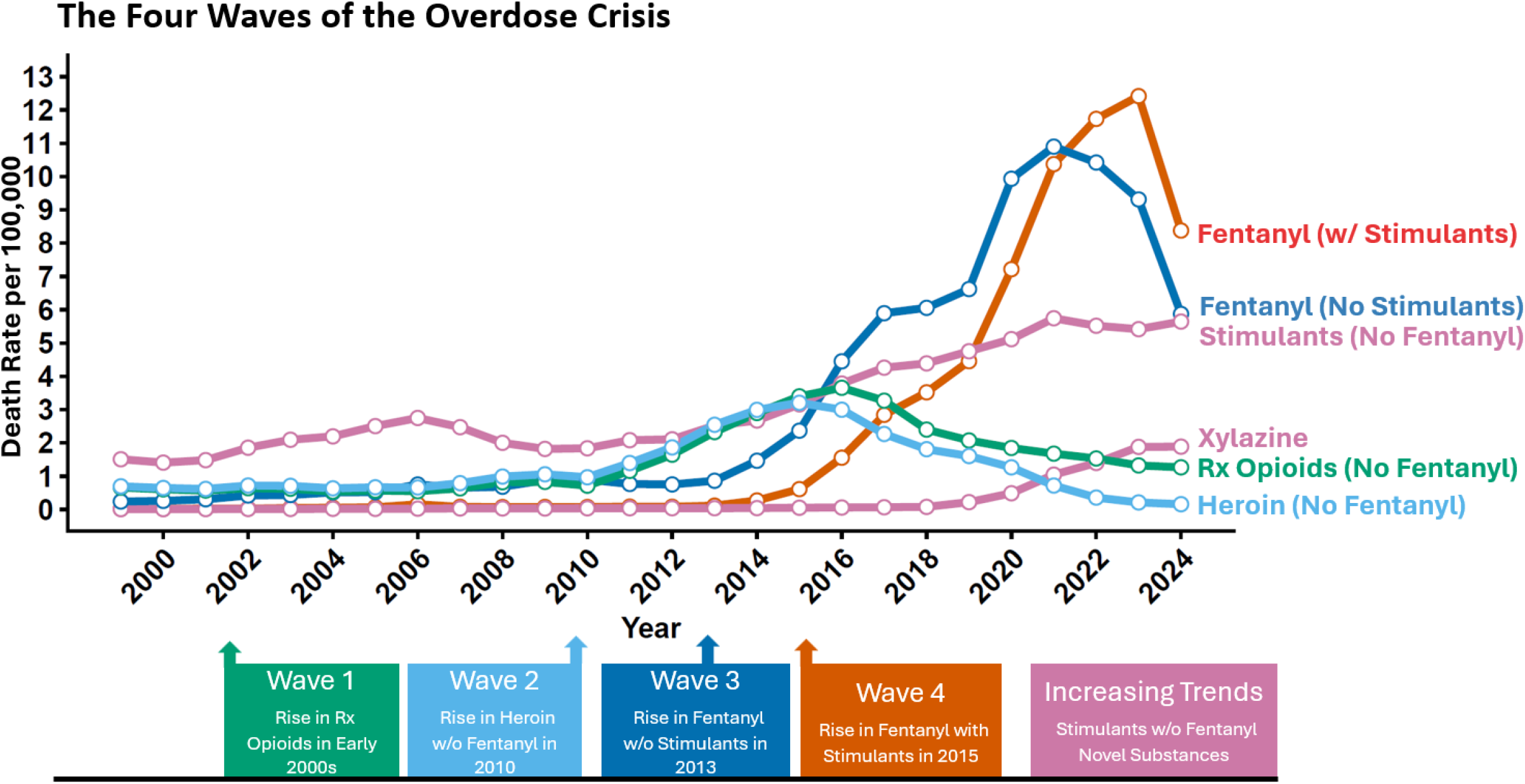
Simplified Representation of the Four Waves of the US Overdose Crisis. Based on the general approach from Friedman and Shover, the four waves of the US overdose crisis are shown in a simplified schema. Wave one is represented by prescription opioids (with fentanyl co-involvement deleted), wave 2 is shown by deaths involving heroin (without fentanyl). Wave 3 and 4 correspond to fentanyl-involved deaths without, and with, stimulants, respectively. Xylazine and stimulants without fentanyl are shown as examples of more recent drug trends that are not decreasing. Note that most xylazine-involved deaths also involve fentanyl. Data from 2024 are provisional and may slightly underestimate final numbers.

The overall decrease in overdose deaths in the US in 2023-2024 has been largely driven by declines in deaths involving illicit fentanyl (Figure 2). Fentanyl-involved deaths without and with stimulants dropped from 31,193 and 41,583 in 2023, respectively, to 19,673 and 28,062 in 2024. In contrast, the number of deaths involving stimulants, without fentanyl, grew from 18,142 in 2023 to 18,907 in 2024, representing 17.3% of all overdose deaths in 2023, and 23.8% of overdose deaths in 2024 (Figure 2).

**Figure 2.**
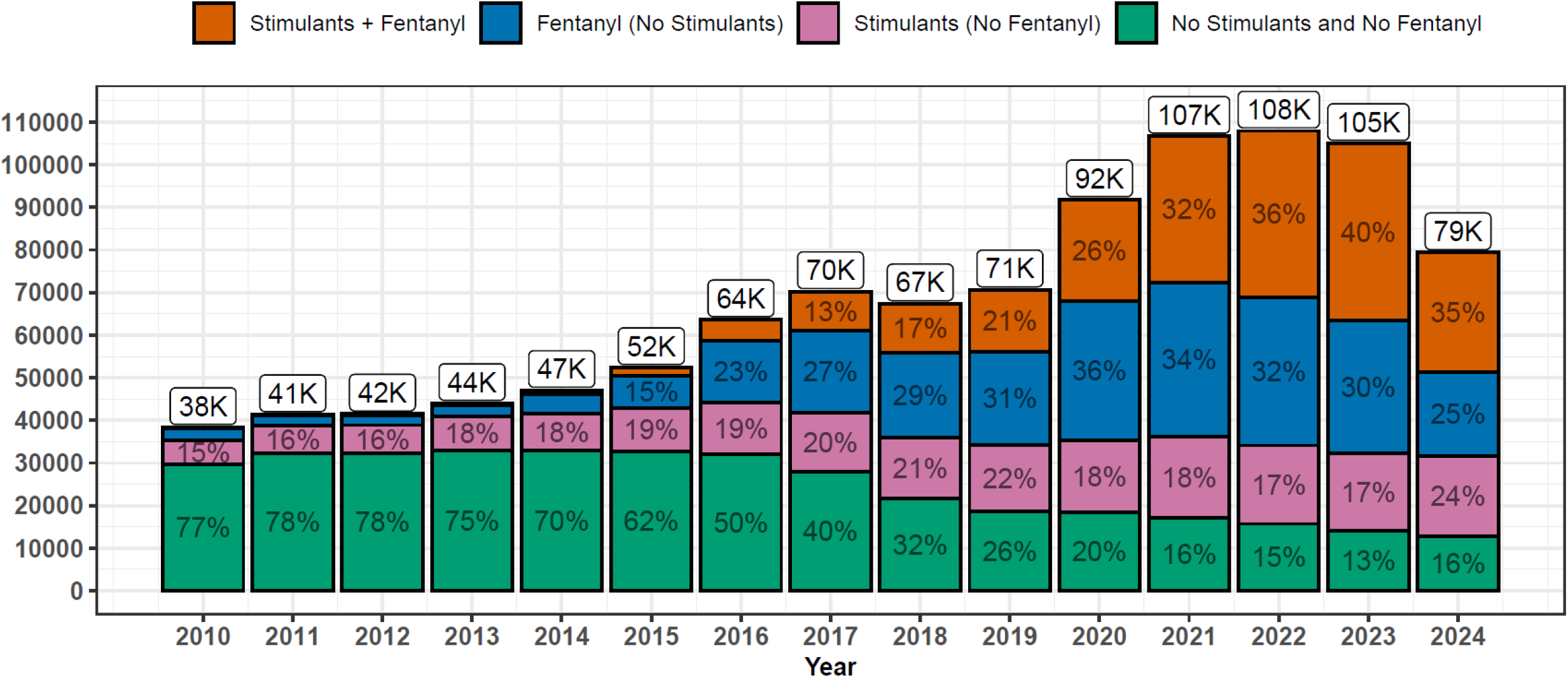
Overdose Deaths by Stimulant and Fentanyl Involvement. The total number of annual overdose deaths each year is shown by categories of fentanyl and stimulant involvement. Data from 2024 are provisional and may slightly underestimate final numbers.

Although all groups saw progress, decreases in overdose death rates also varied by race/ethnicity (Figure 3). Averaging across all racial groups, overdose death rates fell to 23.7 per 100,000 in 2024, representing a 24.4% reduction from 2023. Non-Hispanic Black and African Americans had the largest decrease in overdose death rates between 2023 and 2024 of any group assessed, falling by 14.94 per 100,000 to reach 35.99 per 100,000 (a 29.3% reduction). This death rate nevertheless represented 1.51 times the national average in 2024. Non-Hispanic American Indian and Alaska Native individuals had the highest overdose death rates of any group assessed in 2024, at 50.8 per 100,000 (representing 2.15 times the national average), and experienced an above-average absolute decrease in death rate from 2023 of 12.78 per 100,000, although this represented a below-average relative decrease of 20.1%.

**Figure 3.**
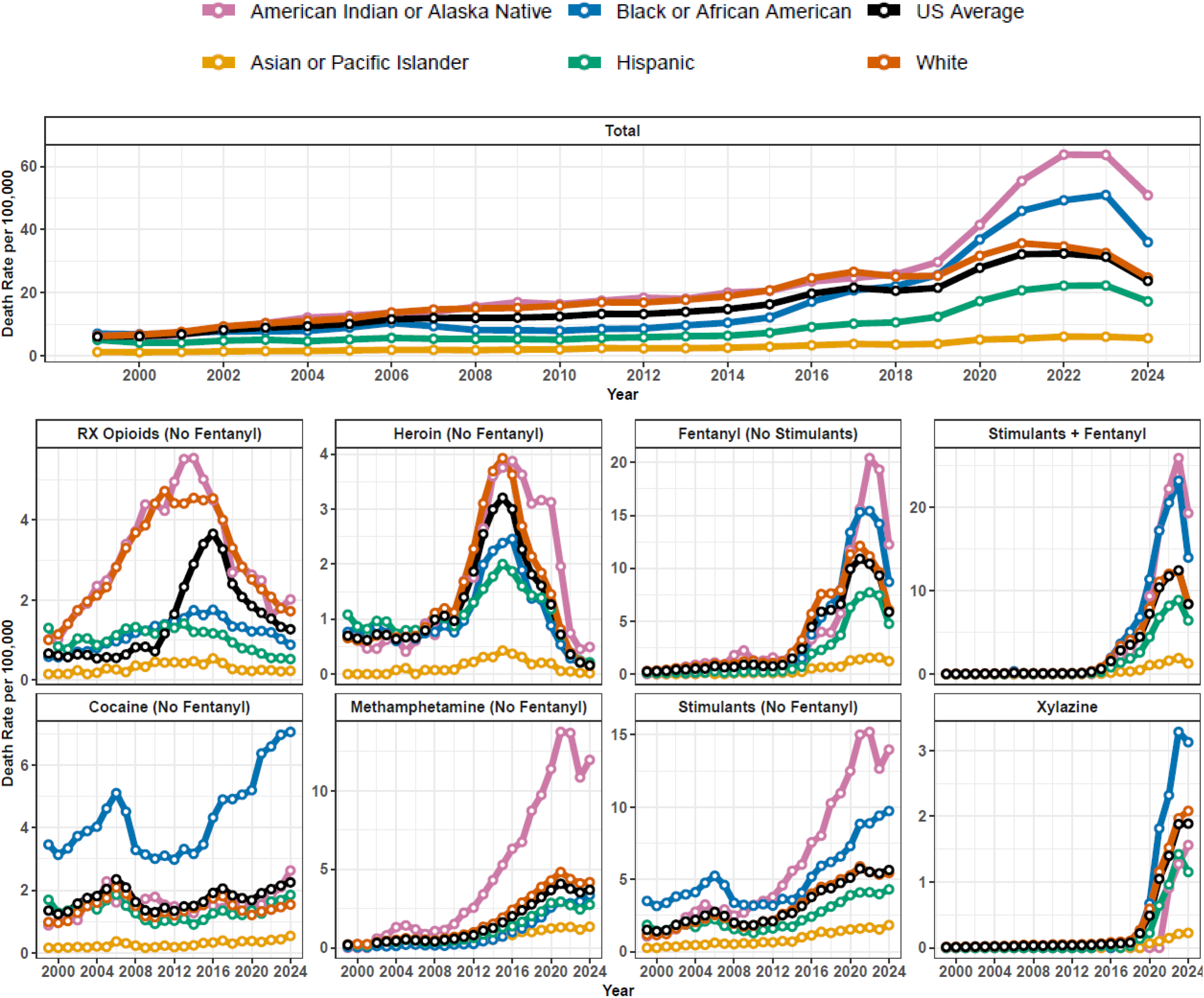
Overdose Deaths by Substance and Race/Ethnicity, 1GGG-2024. Overdose deaths per 100,000 population are shown separately by substance involvement and race/ethnicity between 1999 and 2024. Rx opioids without fentanyl, heroin without fentanyl, fentanyl without stimulants, and fentanyl with stimulants correspond roughly to waves 1,2,3 and 4, of the overdose crisis, respectively. Deaths involving xylazine, and stimulants without fentanyl, represent trends continuing to rise in 2024, despite overall decreases in death totals. Data from 2024 are provisional. Changes in race classification standards within national mortality data around between 2017 and 2018 introduce a potential break in data continuity, although we do not note particularly large discontinuities. Prior to 2018, individuals were all assigned to a single race. Starting in 2018, multiple race was an available category, and these individuals were excluded from the current analysis.

Non-Hispanic White individuals had approximately average overdose death rates in 2024 of 24.8 per 100,000 (1.04 times the national average) and saw a year-over-year percent decrease (23.9%) similar to the national average.

Death rates involving fentanyl with and without stimulants continued to disproportionately affect Non-Hispanic Black and Non-Hispanic American Indian and Alaska Native individuals (Figure 3). Cocaine-involved overdose deaths (with fentanyl deleted) disproportionately affected Non-Hispanic Black and African Americans for all years assessed, with this disparity rising to 3.13 times the national average by 2024. Methamphetamine-involved deaths (with fentanyl deleted) disproportionately affected Non-Hispanic American Indian and Alaska Native individuals, with an overdose rate 3.24 times above the national average in 2024. Xylazine-involved overdose deaths disproportionately affected Black and African Americans, with a rate of 1.66 times the national average in 2024.

## Discussion

After many years of increasing overdose death rates, the US is in the midst of a historic moment in the trajectory of the overdose crisis, with sharply decreasing trends observed between 2023-2024. Although the beginning of these downward trends was previously reported (3), here we provide newfound details documenting their continued trajectory, significance, and profundity. We show that in 2024, for the first time, wave four of the crisis (defined by fentanyl in combination with stimulants), is also decreasing in magnitude. This simultaneous, large-scale reduction in deaths involving fentanyl—both as a standalone driver and in polysubstance combinations—is a remarkable epidemiological shift, accounting for the vast majority of the overall national decline in drug overdose deaths.

Nevertheless, we also highlight that not all segments of the US overdose crisis are shrinking. The persistent increase in drug overdose deaths involving stimulants without fentanyl—even as overall and fentanyl-involved deaths rapidly decline—represents a striking divergence. If these diverging trends continue, stimulants may soon surpass opioids to become the defining addiction-related public health challenge in the US. Other stimulant-related health harms, such as substance-induced psychosis, have also risen (16). It is also important to note that the majority of stimulant-related burden of disease stems from slower epidemiological processes compared to overdose, such as accelerated cardiovascular, neurovascular, pulmonary, and psychiatric consequences (17). As stimulants continue to drive a growing share of addiction-related burden of disease, a focus on overdose deaths as the defining indicator of the public health impact of substance use disorders may be increasingly insufficient. Instead, more comprehensive measures such as disability adjusted life years (DALYS) (17) that account for morbidity may more completely reflect the trajectory of the US addiction crisis.

After several years of rapidly widening inequalities in race/ethnicity—driven by a profound divergence that occurred during the COVID-19 pandemic—2024 showed encouraging decreases in drug overdose death rates across all groups. Most notably, Black and African Americans experienced the largest absolute drop, with rates falling by nearly one-third in a single year, which also contributed to a modest narrowing of racial disparities. Nevertheless, because of the sheer magnitude of prior disparities, inequalities remain large through 2024. In particular, the death rate among American Indian and Alaska Natives remains over double that of the general population. Importantly, the substances with increasing importance through 2024—cocaine, methamphetamine, and xylazine—all show marked racial inequalities, suggesting that disparities may not decrease in future years without intervention. Policies targeting community and system-level factors associated with inequitable health will be needed to address these persistent gaps (13,18,19).

The decreases in overdose deaths in 2023-2024 in the US are likely multifactorial in nature, reflecting several overlapping epidemiological and demographic phenomena (3,20–23). Simulation models have suggested that decreases in the at-risk population may be single most important driver (23), reflecting increased awareness of the risks of illicit fentanyl among the population, as well as extensive premature mortality not replaced by new individuals commencing fentanyl use (3,20,23).

Alternatively other research has suggested that sudden decreases in the availability and/or purity of illicit fentanyl—a “fentanyl drought”—may be a key driving factor, and may better explain the abruptness of the drop in deaths (20,22,24). One analysis used fentanyl purity data from law enforcement seizures, as well as content analysis from social media, to argue that a supply shock occurred in the US and Canada that was temporally aligned with decreases in overdose deaths, and which may have been related to disruptions in availability of fentanyl precursors from China (22).

Several other factors are likely also playing a role. Overdose deaths spiked during COVID-19, and they are now receding alongside several other social, economic, and epidemiological consequences of the pandemic (25). The availability and use of the opioid receptor antagonist naloxone, which is a short-term reversal agent for opioid overdoses, have also been considerably scaled up in recent years (26,27). The widespread shift from injecting to smoking opioids has been associated with decreased overdose risk (28,29). Further research is needed to assess the relative contributions of these factors.

Some of these observed trends may also simply reflect, at least in part, an overall ‘regression to the mean’ as trends homogenize in the context of nearly universal decreases. Examining demographic categories highlights that the groups with the highest drug overdose death rates—such as Black and African American, and American Indian and Alaska Native populations—generally experienced the greatest absolute decreases, which would align with this statistical phenomenon.

### Limitations

Death records from 2024 are provisional and may change when final records are released (30). The race/ethnicity categorizations used here have limitations, especially for American Indian and Alaska Native individuals, which may mask the magnitude of inequalities (31). Furthermore, changes in race classification standards within national death data between 2017 and 2018 introduce a potential break in data continuity (32). Multiracial individuals were excluded from both death and population counts starting in 2018 in our analysis, which represents a limitation that may subtly affect longitudinal estimates. Nevertheless, trends analyzed between 2018 and 2024 are internally consistent.

## Conclusions

Recent decreases in overdose deaths are encouraging and relatively unprecedented. Despite recent progress, US overdose death rates remain a global outlier (33). The country saw about 80,000 deaths in 2024. If the US population had the same overdose death rate as Western Europe, about 9,000 lives would have been lost (17). Greater investments are needed to sustainably reverse the course of this large-magnitude, preventable death crisis, via evidence-based approaches such as medications for opioid use disorder, harm reduction, and access to addiction-related treatment.

Some encouraging, yet early, improvements in racial/ethnic inequalities in overdose death rates have also been seen, after profoundly worsening during the COVID-19 pandemic (13). Yet disparities remain, even as the national trend in deaths now declines. More progress can be made with tailored solutions to reduce overdose deaths among the most affected groups (13).

The fraction of drug overdose deaths involving stimulants without fentanyl and those involving xylazine continue to increase, highlighting shifting challenges in the overdose crisis.

## Data Availability

All data are publicly available through the US CDC WONDER platform.

